# A novel food frequency questionnaire for Brazilian adults based on the Nova classification system: development, reproducibility and validation

**DOI:** 10.1101/2024.04.19.24305963

**Authors:** Evelyn Oliveira da Silva Frade, Kamila Tiemann Gabe, Caroline dos Santos Costa, Daniela Neri, Euridice Martinez Steele, Fernanda Rauber, Josiane Steluti, Renata Bertazzi Levy, Maria Laura da Costa Louzada

**Author notes:** Provide full correspondence details here including e-mail for the Corresponding author: Evelyn Oliveira da Silva Frade | | Centre for Epidemiological Research in Nutrition and Health, School of Public Health, University of São Paulo.

## Abstract

The Nova system categorizes foods according to processing levels, and dietary intake instruments not developed to assess this criteria may introduce bias in epidemiological studies. To address this gap, we developed and validated the Nova Food Frequency Questionnaire (NovaFFQ) for Brazilian adults. The NovaFFQ includes commonly consumed foods and drinks based on 2017-2018 National Food Survey data. Reproducibility was assessed by comparing NovaFFQ estimates on two occasions. Criterion validity was assessed by comparing the mean dietary contribution of Nova groups obtained from the first NovaFFQ against two Nova24h. Strong reproducibility was observed with an intraclass correlation coefficient (ICC) of 0.91 for all Nova groups. Criterion validity showed a moderate ICC, ranging from 0.61 to 0.65, and substantial agreement in ranking individuals, as indicated by prevalence and bias-adjusted kappa, ranging from 0.70 to 0.74. The NovaFFQ is a valid instrument for assessing food consumption according to food processing.

## Introduction

The Nova classification system classifies foods based on the degree and purpose of industrial processing. It divides all foods into four distinct groups: (1) unprocessed or minimally processed foods (e.g., fruits, vegetables, meat), (2) processed culinary ingredients (e.g., sugar, salt, oil), (3) processed foods (e.g., jam, cheese), and (4) ultra-processed foods (UPF) (e.g., crackers, soft drinks, ready-to-heat or ready-to-eat meals) (Monteiro et al. 2019).

Nova has been widely used for understanding the impacts of the degree of processing of dietary patterns on health and food systems worldwide. Studies have shown, for example, a decline over time in sales and consumption of unprocessed and minimally processed foods and processed culinary ingredients, and an increase in ultra-processed foods globally (Baker et al. 2020). Hundreds of studies have documented the effects of food processing on human health. A high consumption of UPF has been associated with a worse dietary nutritional profile in several countries (Martini et al. 2021; Martinez Steele et al. 2022) and a greater risk of weight gain and several non-communicable chronic diseases such as type 2 diabetes, hypertension, and some cancers (Delpino et al. 2022; Askari et al. 2020; Wang et al. 2022).

Despite all this evidence, a frequent limitation of the above-mentioned studies is the use of instruments that were not developed and validated to estimate food intake according to the Nova system. Current dietary assessment instruments do not probe respondents for the level of detail researchers need to make accurate Nova classifications.

Traditional 24-hour recalls, which provide food-level information, often lack needed detail (e.g., whether foods are prepared at home using conventional cooking methods vs. pre-prepared/packaged, or brand names). To overcome the limitation of the 24-hour recall, researchers from the Centre for Epidemiological Studies in Nutrition and Health at the University of São Paulo (NUPENS/USP) developed a 24-hour food recall specifically designed to assess food consumption according to the Nova system. The Nova24h is a web-based self-completed instrument that assesses foods and drinks consumed over the last 24 hours. It showed good performance when compared to a traditional 24-hour recall applied by an interviewer to capture the energy contribution of each Nova group and to classify individuals according to quintiles of each Nova food group consumption (Neri et al. 2023).

FFQs are other instruments widely used for dietary assessment in epidemiological studies. These instruments are more easily administered than 24-hour recalls, they capture intake over a long period of time and may better estimate usual dietary intake with a single application (Cade et al. 2001; Marchioni, Gorgulho, and Steluti 2019). Large prospective studies have used FFQs to assess long-term health effects of food processing (Canhada et al. 2020; Hang et al. 2023; Rico-Campà et al. 2019). However, food misclassification is a particular concern for food-frequency questionnaires. Their closed food list may not include all necessary details to classify the items into Nova groups, and they may also mix home-prepared and ultra-processed foods in the same item. For example, studies using these instruments may misclassify a packaged cake as a culinary preparation made from unprocessed and minimally processed foods and processed culinary ingredients instead of as ultra-processed (Touvier et al. 2023).

To the best of our knowledge, only three FFQs have been previously designed for estimating food consumption according to Nova classification (Motta et al. 2021; Amorim, Prado, and Guimarães 2020; Dinu et al. 2021). Motta and colleagues developed an FFQ for Brazilian children from the Midwest, and Amorim, Prado, and Guimarães developed an FFQ for Brazilian adults from the Northeast. However, none of these instruments have been validated yet. Conversely, Dinu and colleagues adapted and validated an FFQ for Italian adults, demonstrating good test-retest reliability and moderate to good validity.

Given that no instruments were explicitly designed and validated to assess the intake of the Nova groups across the entire Brazilian adult population, this study aimed to describe the development and evaluate reproducibility and validity of a Food Frequency Questionnaire specifically designed to estimate food consumption in line with the Nova classification in Brazilian adults.

## Materials and methods

### 1. Development of NovaFFQ

The Nova Food Frequency Questionnaire (NovaFFQ) is a web-based, self-completed, quantitative instrument for the past 12 months. We developed the NovaFFQ in nine steps using data from 24-hour recalls of adults from the 2017-2018 National Food Survey (POF 2017-2018) (IBGE, 2020). The development of the instrument is summarised in Figure 1 and detailed below.

1. We grouped identical foods that were coded differently in the database (e.g. “mandioca” and “aipim” are different names and codes for cassava) or that were prepared in various ways (e.g. roast meat or grilled meat).
2. We estimated the dietary contribution of each food to total energy intake.
3. We included in a food list all foods accounting for 95% of the calories consumed by Brazilian adults.
4. From the compiled food list, we identified and differentiated foods that could be classified into alternative Nova groups, such as: breads or cakes prepared from scratch (with flour and other ingredients) at home, or at a restaurant or bakery, and ultra-processed versions of breads and cakes. Each of these items were replaced by two separate food items from the alternative Nova groups, each including a description of all relevant information for their accurate identification and subsequent Nova classification. For instance, for lasagna two different items were created: (1) “lasagna prepared at home or at a restaurant using conventional cooking methods” and (2) “ready-to-heat lasagna”. Likewise, cake was described as (1) “homemade or bakery cake” and (2) “store-bought, prepacked, branded cake or prepared from a packed mix”.
5. We include items that are usually added to foods at the time of consumption, such as sugar, butter, and sauces.
6. For each item, we established the standardised portion as the most frequently consumed portion (e.g. for rice, the standardised portion was “1 serving spoon”), based on data from POF 2017-2018.
7. We defined the response options for frequency of consumption and usual amount consumed based on those used by a previously validated FFQ designed to estimate the weight contribution of Nova food groups in Italian adults (Dinu et al. 2021). The options for frequency were: “Never or rarely”, “1 time per month”, “2-3 times per month “, “1 time a week”, “2 times a week”, “3 times a week”, “4 times a week”, “5 times a week”, “6 times a week”, “Daily”. The options for the usual amount were: “0.5”; “1.0”; “1.5”; “2.0”, “2.5”; “3.0”, “+3.5”.

**Figure 1.**
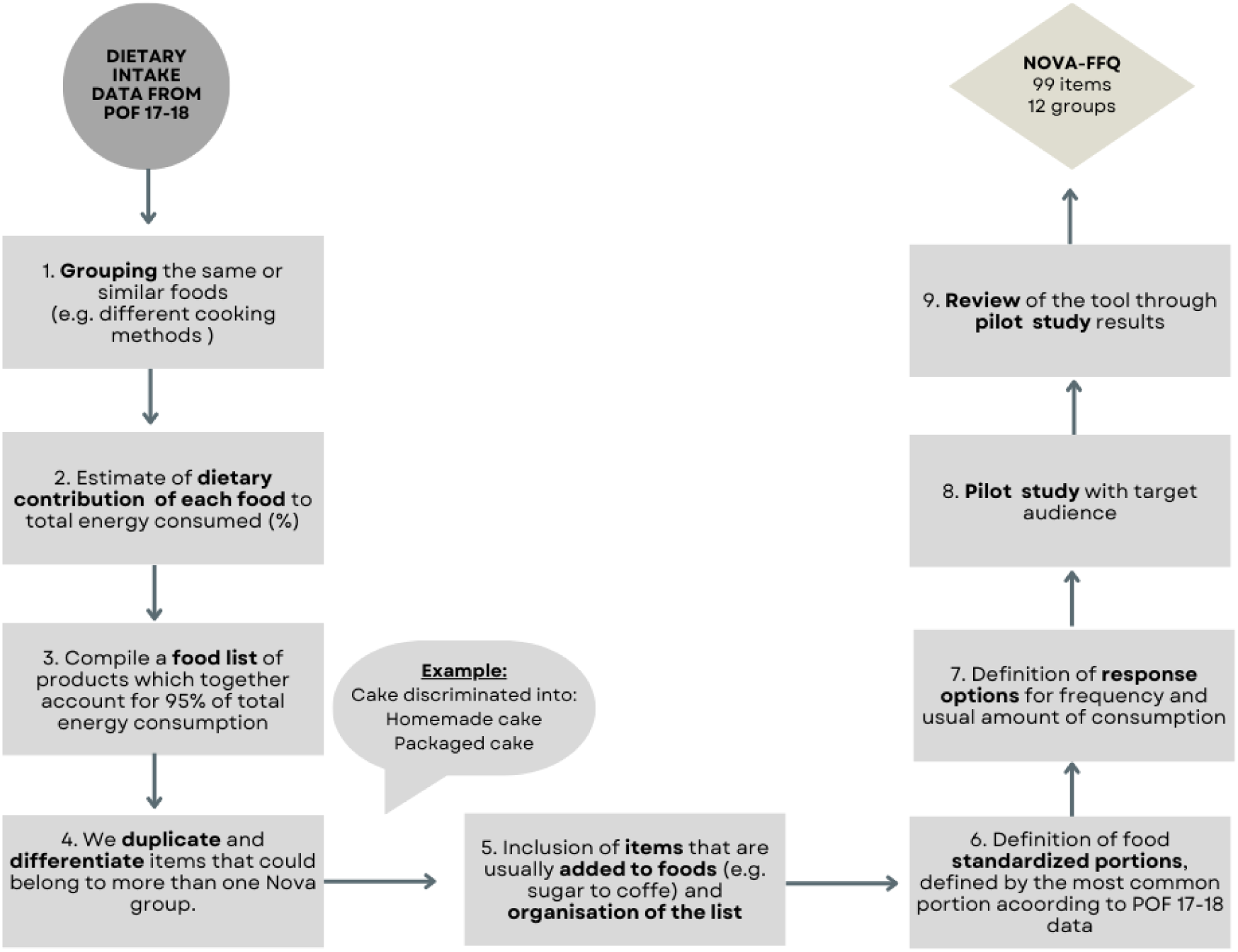
Flowchart of the development and pilot study of the NovaFFQ.

After that, experts from the NUPENS/USP with experience in analysing food consumption according to Nova were invited to review the instrument. The experts analysed the suitability of the food list (after step 5) and standardised portions (after step 6) and suggested adjustments. The initial list of foods obtained from POF’s data in step 3 contained 62 items. After incorporating details to ensure accurate classification into the Nova groups, the number of items of the initial version of the questionnaire corresponded to 111 (step 5). The NovaFFQ includes items within twelve sections in the following order: “1. Cereals and pasta”; “2. Beans”; “3. Hamburgers, meats, and eggs”; “4. Vegetables”; “5. Roots and tubers”; “6. Fruits”; “7. Cakes, pastries, desserts, and breakfast cereals”; “8. Breads, biscuits, snacks, and pizzas”; “9. Processed meat and cheese”; “10. Drinks”; “11. Nuts”; and “12. Items added to foods or preparations”. Respondents are provided with brief instructions on how to complete the NovaFFQ. Then, for each food item included in the questionnaire, participants are asked two questions: a) frequency of consumption; and b) usual amount consumed based on the standardised portion.

### 1.1 Pilot study

Next, we conducted a pilot study to verify the feasibility and interpretability of the NovaFFQ in a convenience sample of 20 adults aged 18 years or older, of both sexes, residing in Brazil (depicted in steps 8 and 9 from Figure 1). We excluded pregnant or lactating women, nutrition undergraduate students, and dietitians.

We recruited participants through social networks with posts on the NutriNet-Brasil study and NUPENS account on Instagram and Twitter. We had 79 applications, from which we selected 20 participants, aiming for the greatest possible diversity in terms of sex, macro-regions of residence, age and schooling. After the selected participants had filled out the consent form and the NovaFFQ, we conducted an online interview to capture participants’ understanding regarding initial instructions, response options for frequency and portions, standardised portions, and descriptions of food items. We tabulated the data from each interview, and two researchers analysed and discussed the data. Some modifications were carried out, mainly in the section names, the description of items, the examples, and the grouping of similar items. For example, the juice item initially described as ‘Natural fruit juice (fresh or pasteurised)’, was simplified to ‘Natural fruit juice’, after the pilot study, because the term ‘pasteurised’ was not clear. After adjustments, the final number of items of NovaFFQ was 99, and the average time to complete the NovaFFQ during the pilot study was 25 minutes. Supplementary material 1 provide the NovaFFQ in English (free translation).

### 2. Reproducibility analysis and criterion validation

#### 2.1 Study participants and data collection

We conducted the NovaFFQ validation in a subsample of the ongoing NutriNet-Brasil study launched in January 2020. The NutriNet-Brasil study aims to prospectively investigate the relationship between dietary patterns and morbidity and mortality from noncommunicable diseases in Brazil. The cohort includes individuals aged 18 years or older, with internet access, and residing in Brazil.

Every six months, participants of the NutriNet-Brasil study respond to the Nova24h recall, which was specifically developed and validated to estimate food consumption based on industrial processing (NERI et al., 2023).

The Nova24h recall is a self-reported and web-based 24-hour recall. Participants are asked 57 key questions, and then, when they answer “yes” to one of them, are presented with additional questions about the type of food (e.g. “homemade bread”), amount consumed (e.g. “1 slice”), and other details (e.g. “whole grain bread”). All these consist of 395 close-ended questions. The Nova24h system provides a database with all foods and drinks consumed, as well as the nutritional composition, using the Brazilian Food Composition Table 7.0 (TBCA), and the Nova classification for each item. Within this dataset, mixed-dishes are broken down into their individual ingredients utilising a TBCA recipe-database. Further details about Nova24h can be found in Neri et al. (2023). The Nova24h was the reference instrument in the current study.

To validate the dietary intake estimated by NovaFFQ, data from two Nova24h recall was considered. Considering the five macro-regions of Brazil and a sample size of at least 50 individuals for each, we defined an intended sample size of 300 individuals for reproducibility and validation (Cade et al. 2001). Also, considering the observed refusals to respond to additional questionnaires and withdrawals from the NutriNet-Brasil study, as well as possible energy outliers reports on Nova24h, we invited 1,200 participants, who had completed two Nova24h recalls, within the past 12 months. Exclusion criteria were being pregnant or breastfeeding women and/or being nutrition undergraduate students – and dieticians.

Participants were informed about the study procedures and completed the informed consent form. Then, they were asked to complete the NovaFFQ on two different occasions, over a period of four to six weeks between administrations. All procedures in this study were approved by the Research Ethics Committee of the School of Public Health of the University of São Paulo (approval number: #4.795.478).

#### 2.2 Data processing

The respective portions of each food reported in NovaFFQ were converted into grams and, thereafter, energy using TBCA 7.0. Mixed-dishes were disaggregated into their ingredients (e.g., home-prepared beans were broken down into beans, oil, garlic, and salt) using standardised recipes from TBCA 7.0. The same criteria previously developed and validated to classify Nova24h food items according to Nova (Neri et al. 2023) were applied to the NovaFFQ.

The estimated daily energy consumed from each food reported in the NovaFFQ was estimated by the following equation:

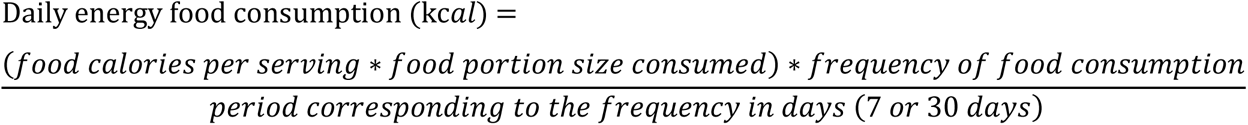

NovaFFQ items that were reported in a grouped form (e.g., rice, including white rice and brown rice) had their energy weighted for each food according to the proportion of consumption of the Brazilian population.

#### 2.3 Statistical analysis

We described sample characteristics with mean and standard deviation for age, and frequency distribution for sex (male, female), macro-region of residence (North, Northeast, Centre-West, Southeast, South), and level of education (Less than elementary, Elementary, Secondary, Completed college/university). To compare the instruments, we estimated the dietary contribution of energy from each Nova group as percentage of total energy intake (%). For the Nova24h recall, we estimated the mean between the two measurements.

Outliers for total energy intake estimated by NovaFFQ or Nova24h were excluded from the analysis according to the following criteria: for males, energy intake below 800 calories and above 4,000 calories; for females, energy intake below 500 calories and above 3,500 calories (Marchioni, Gorgulho, and Steluti 2019).

To evaluate the reproducibility of the instrument, the test-retest method was used. The reproducibility study sample consisted of participants who had two valid NovaFFQ assessments. We compared the dietary contribution for the total energy intake of Nova’s groups in the first and second applications of NovaFFQ. We estimated the intraclass correlation coefficient (ICC) and 95% confidence interval using the two-way mixed effects model. In the reproducibility analysis, ICC measures the degree of agreement between the individuals’ measurements taken at separate times. Values lower than 0.5 indicate poor agreement, values between 0.5 and 0.75 indicate moderate agreement, values between 0.75 and 0.90 indicate good agreement and values above 0.90 indicate excellent agreement (Koo and Li 2016).

To assess the criterion validity, we compared the energy contribution of Nova’s groups obtained in the first application of the NovaFFQ against the mean estimates obtained in the two Nova24h recalls. The validation study sample was composed by participants with the first valid NovaFFQ assessment and two valid Nova24h assessments. We estimated the ICC and 95% confidence intervals using the two-way mixed effects model to assess the degree of agreement between the methods. As in reproducibility analysis the coefficient measures the degree of agreement between the individuals’ measurements made by different instruments.

We divided the sample into quintiles of energy contribution of each Nova group using both methods (Nova24h and NovaFFQ) to assess the ability of the NovaFFQ to rank individuals according to the level of consumption of each Nova group. We assessed the proportion of participants who were correctly classified (same quintile), correctly or adjacently classified (same or next quintile), and grossly misclassified (highest quintile by NovaFFQ and lowest by Nova24h, or vice versa). We also estimated the prevalence-adjusted and bias-adjusted kappa (PABAK) to assess the agreement of sample classification into quintiles. For PABAK, values between 0.00 and 0.20 indicate low agreement, between 0.21 and 0.40 indicate acceptable agreement, values between 0.41 and 0.60 indicate moderate agreement, values between 0.61 and 0.80 indicate substantial agreement, and values above 0.8 indicate almost perfect agreement (Landis and Koch 1977). Analyses were performed using Stata version 17.0 and R Studio software.

## Results

### Study participants

A total of 1,200 participants were invited to participate in the study, of whom 409 completed the first NovaFFQ. After excluding 32 individuals due to outlier reports for total energy intake in the Nova24h recalls and the first NovaFFQ, we had a final sample of 377 participants for the validity analysis. Out of the 377 participants, 248 completed the second NovaFFQ. Five participants were excluded due to outlier reports for total energy intake, resulting in a sample size of 243 for the reproducibility analysis (Figure 2).

**Figure 2.**
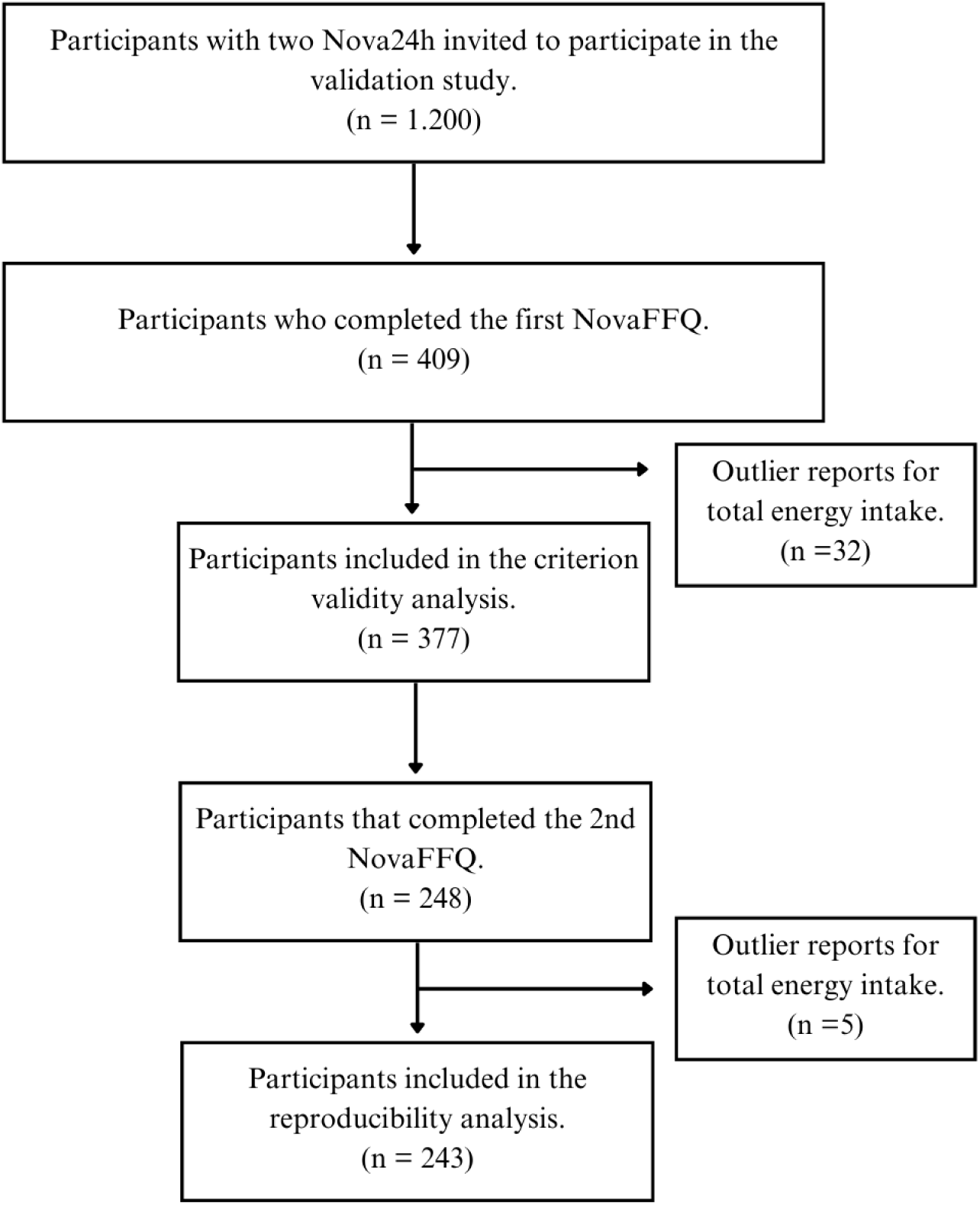
Flowchart of criterion validity and reproducibility analysis samples.

Table 1 presents the sociodemographic characteristics of both the reproducibility and criterion validation samples. In the reproducibility sample, the mean age was 45.6±12.3 years, and 55.6% were female. About one third of the participants resided in the Southeast region of Brazil (33.8%) and over three fourths had completed college/university (77.4%). In the criterion validation analysis sample, the mean age was 44.1±12.7 years and 55.2% were female. About one third of the participants lived in the Southeast region of Brazil (31.3%) and about three fourths had completed college/university (73.5%).

**Table 1.** Socio-demographic characteristics of study participants.

| Characteristics | Reproducibility study<br>(n=243) |  | Validation criterion study<br>(n=377) |  |
| --- | --- | --- | --- | --- |
| | Mean ± SE | n (%) | Mean ± SE | n (%) |
| Age (years) | 45.6 ± 12.3 | | 44.1 ± 12.7 | |
| *Sex* |  |  |  |  |
| Female |  | 135 (55.6) |  | 208 (55.2) |
| Male |  | 108 (44.4) |  | 169 (44.8) |
| *Region* |  |  |  |  |
| North |  | 28 (11.5) |  | 48 (12.7) |
| Northeast |  | 50 (20.6) |  | 79 (21.0) |
| Centre-West |  | 46 (18.9) |  | 69 (18.3) |
| Southeast |  | 82 (33.8) |  | 118 (31.3) |
| South |  | 37 (15.2) |  | 63 (16.7) |
| *Educational level* |  |  |  |  |
| Less than elementary |  | 9 (3.7) |  | 14 (3.7) |
| Elementary |  | 6 (2.5) |  | 12 (3.2) |
| Secondary |  | 40 (16.5) |  | 74 (19.6) |
| Completed college/university |  | 188 (77.4) |  | 277 (73.5) |

### Reproducibility analysis

The dietary contributions of Nova groups were similar between NovaFFQ administrations. Unprocessed and minimally processed food showed a mean absolute difference of 0.40 percentage points (pp), the processed culinary ingredients showed a difference of 0.10 pp, the processed foods a difference of 0.11 pp, and the ultra-processed group a difference of −0.61 pp. Additionally, we observed excellent agreement, with an ICC of 0.91 for all Nova groups, indicating that the NovaFFQ demonstrated a good ability to produce consistent results over time (Table 2).

**Table 2.** Dietary contribution (% of total energy intake) of Nova groups using Nova Food Frequency Questionnaire applied on two different occasions. Reproducibility study. (n=243)

| Nova groups | % of total energy intake |  |  |  | Mean difference<sup>1</sup> | ICC<sup>2</sup> | 95% IC |  |
| --- | --- | --- | --- | --- | --- | --- | --- | --- |
|  | NovaFFQ 1 |  | NovaFFQ 2 |  |  |  |  |  |
|  | Mean | SE | Mean | SE |  |  |  |  |
| Unprocessed and minimally processed foods | 55.7 | 0.7 | 55.3 | 0.7 | 0.40 | 0.91 | 0.88 | 0.93 |
| Processed culinary ingredients | 11.7 | 0.3 | 11.6 | 0.3 | 0.10 | 0.91 | 0.88 | 0.93 |
| Processed foods | 17.3 | 0.6 | 17.2 | 0.6 | 0.11 | 0.91 | 0.89 | 0.93 |
| Ultra-processed foods | 15.3 | 0.6 | 15.9 | 0.6 | -0.61 | 0.91 | 0.88 | 0.93 |

<sup>1</sup>Absolute difference between the first and second administration
<sup>2</sup>Intraclass correlation coefficients

### Criterion validation analysis

The comparison of the dietary contribution for unprocessed and minimally processed foods revealed a mean absolute difference of 5.96 pp between the estimate of the NovaFFQ and the reference instrument (mean of two Nova24h recalls). For processed culinary ingredients, the difference was 0.34 pp, while for processed and ultra-processed foods, it was −1.88 pp and −4.42 pp, respectively. We observed moderate agreement between the instruments, as indicated by the ICC ranging from 0.61 for processed and ultra-processed foods to 0.65 for unprocessed and minimally processed foods (Table 3).

**Table 3.** Dietary contribution (% of total energy intake) of Nova groups using the mean of two Nova24h and the first Nova Food Frequency Questionnaire. Criterion validation study. (n=377)

| Nova groups | % of total energy intake |  |  |  | Mean difference<sup>1</sup> | ICC<sup>2</sup> | 95% IC |  |
| --- | --- | --- | --- | --- | --- | --- | --- | --- |
|  | Nova24h |  | NovaFFQ |  |  |  |  |  |
|  | Mean | SE | Mean | SE |  |  |  |  |
| Unprocessed and minimally processed foods | 50.4 | 0.7 | 56.2 | 0.6 | 5.96 | 0.65 | 0.48 | 0.76 |
| Processed culinary ingredients | 11.3 | 0.3 | 11.7 | 0.3 | 0.34 | 0.63 | 0.54 | 0.70 |
| Processed foods | 18.5 | 0.6 | 16.7 | 0.5 | -1.88 | 0.61 | 0.52 | 0.68 |
| Ultra-processed foods | 19.8 | 0.6 | 15.4 | 0.5 | -4.42 | 0.61 | 0.47 | 0.71 |

<sup>1</sup>Absolute difference between the first NovaFFQ and Nova24h
<sup>2</sup>Intraclass correlation coefficient

Table 4 presents the distribution of the sample into quintiles of dietary contribution of each Nova group estimated by NovaFFQ and the reference instrument, with the percentage of agreement in each quintile and the PABAK statistic. Overall, we observed percentages higher than 67% of correctly or adjacent classification and percentage lower than 15% of gross misclassification for all Nova groups. We also observed a higher percentage of agreement in the lowest (Q1) and the highest (Q5) quintiles of consumption. PABAK estimates ranged between 0.70 and 0.74, indicating substantial agreement between the instruments in ranking individuals into quintiles.

**Table 4.**
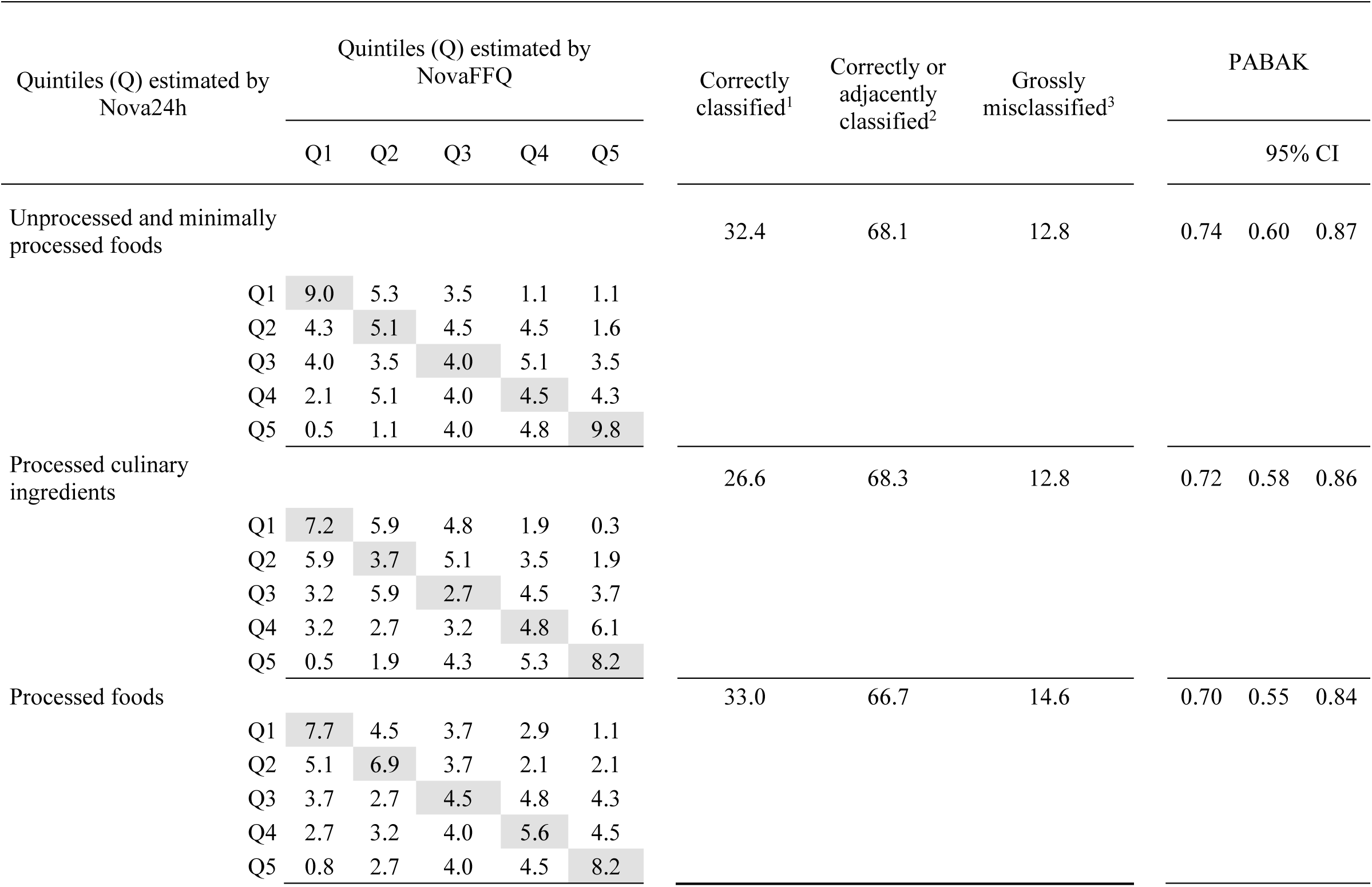
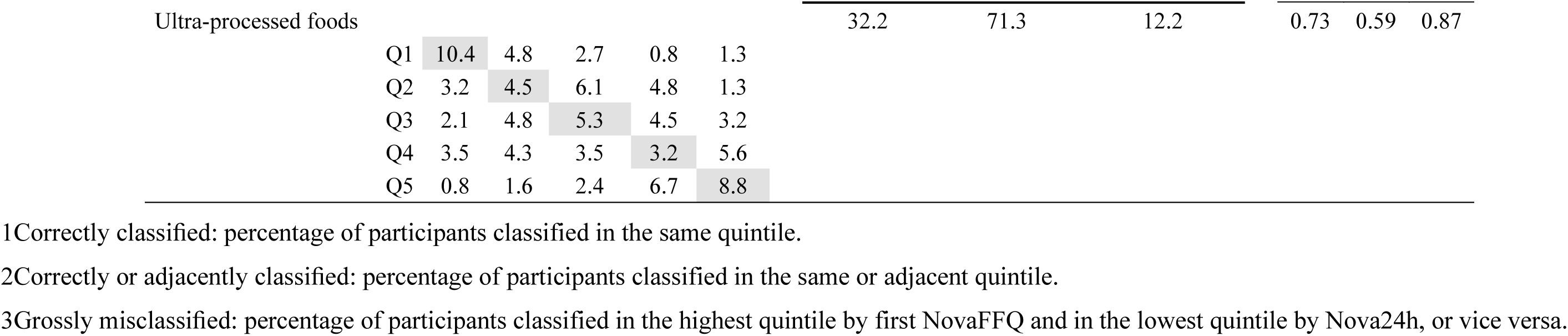
Agreement and cross-classification between participants classification according to quintiles of the dietary energy contribution of each Nova group estimated by the mean of two Nova24h and the first Nova Food Frequency Questionnaire. (n=377)

Supplementary material 2 presents the mean dietary contribution of Nova subgroups estimated by the reference instrument and the NovaFFQ, as well as the difference between these estimates and the ICC of each subgroup. The largest difference between the instruments was observed for the unprocessed and minimally processed foods group, and fruits were responsible for almost 50% of that difference (mean difference of 2.9 pp).

## Discussion

This study describes the development and evaluation of reproducibility and validity of a food frequency questionnaire designed to assess food consumption based on the Nova classification in the adult Brazilian population. The questionnaire underwent a rigorous review by experts in Nova classification and dietary assessment and was tested in a pilot study with Brazilian adults. Results demonstrated a strong ability to replicate energy estimates from Nova groups consistently over time and moderate criterion validity to estimate food consumption according to the Nova system. The instrument also exhibited significant validity in ranking individuals according to their level of consumption into the four Nova groups.

The NovaFFQ is the third validated instrument developed to assess food consumption based on the degree of processing in the Brazilian population, together with the Nova24h recall (Neri et al. 2023) and the Nova Screener (Costa et al. 2023). The NovaFFQ is a low-cost questionnaire that can be administered repeatedly over time and, as other FFQs, may be particularly valuable for epidemiological studies aiming to assess the long-latency effects of an exposure (e.g., consumption of ultra-processed foods) on outcomes such as cancer (Kac, Sichieri, and Gigante 2007). It may also be useful for assessing food consumption before an event that might modify food consumption, such as usual dietary intake prior to pregnancy. The significant distinction of NovaFFQ is its capacity to assess food consumption according to food processing, providing immediate estimates of usual consumption within Nova’s four food groups.

To the best of our knowledge, only three FFQs have been specifically developed to assess food consumption according to the degree of processing. Dinu and colleagues (2021) adapted a pre-existing FFQ developed for the Italian adult population by incorporating information on food processing into the instrument. This FFQ was validated by comparing the FFQ dietary contribution of each Nova group expressed in percentage of grams per day against the weighted seven-day dietary record mean contributions. They obtained a good ICC ranging from 0.77 to 0.85, like the moderate ICC obtained in the NovaFFQ (Dinu et al. 2021). The other two FFQs were developed but not validated to assess dietary intake according to Nova among Brazilians from specific regions, one for adults in the Northeast and the other for children in the Midwest (Motta et al. 2021; Amorim, Prado, and Guimarães 2020).

The validation analysis of the NovaFFQ indicates satisfactory agreements. The differences between means of unprocessed/minimally processed foods may be mainly attributed to the overestimation of fruits consumption in the NovaFFQ when compared to Nova24h. One plausible explanation for this discrepancy could be the seasonality of fruit consumption. Since some fruits are available only during specific periods of the year, respondents may provide overestimated measures in the NovaFFQ without considering that the frequency of consumption might have varied throughout the previous year.

Another explanation refers to the trait of social desirability, which is an individual’s tendency to give adequate responses to social norms to avoid criticism (Hebert et al. 1997). The FFQs may be particularly more affected by social desirability, since they suffer greater influence of individual’s perception on their own food consumption (Willet 1998). Some studies have shown a significant effect of social desirability on self-reported estimates of food consumption. The importance of regular fruit consumption for a healthy diet is increasingly known, which, associated with the trait of social desirability, may have influenced the overestimated response of participants to foods considered healthy. Two studies, in different populations, assessed the effect of social desirability (measured by a validated scale) specifically on fruit and vegetable consumption. They found a positive association between scores on the scale and frequency and quantity of fruit and vegetable consumption (Barros, Moreira, and Oliveira 2005; Di Noia, Cullen, and Monica 2016).

One of the most significant findings of the present study was the substantial agreement of the NovaFFQ to rank individuals according to the level of consumption of the four Nova groups, allowing the differentiation of high and low consumers of each group. This is particularly valuable as most prospective studies on diet and disease incidence examine associations by comparing disease risk across categories of the dietary factor of interest. More recently, the degree of processing has been extensively considered, and studies have been dividing participants into three to five categories, always based on their level of consumption of ultra-processed foods. Individuals in the lowest consumption category are used as a reference in these studies. For instance, a meta-analysis of 23 studies showed that the highest category of ultra-processed food consumption was associated with a 25% and 34% increased risk of cardiovascular and cerebrovascular diseases, respectively (Pagliai et al. 2020).

Previous cohort studies assessing the effect of UPF consumption derived from FFQ on health have often highlighted as a limitation the use of an FFQ not specifically designed to assess degree of processing (to obtain UPF estimates). For instance, Hang et al. (2023) investigated, in the Nurses’ Health Study, Nurses’ Health Study II, and Health Professionals Follow-up Study, the consumption of ultra-processed foods and the risk of colorectal cancer precursors by comparing the risk between the first quintile of consumption (lowest) and the subsequent ones and the cohort from the University of Navarra (SUN, from the Spanish Seguimiento Universidad de Navarra) analysed the consumption of ultra-processed foods and all-cause mortality by comparing mortality between quartiles of consumption of ultra-processed foods, using the first quartile as a reference (Rico-Campà et al. 2019). In Brazil, the Longitudinal Study of Adult Health (ELSA-Brazil, from the Portuguese Estudo Longitudinal de Saúde do Adulto-Brasil) evaluated the consumption of ultra-processed foods and the risk of overweight and obesity by comparing the risk between the first and fourth quartiles of consumption of ultra-processed foods (Canhada et al. 2020). The results of these studies could be improved by removing this limitation. This highlights the relevance of the now validated NovaFFQ.

This study has limitations and strengths. Strengths include the use of data from a nationally representative survey of the Brazilian population, POF 17-18, which allowed us to incorporate the foods consumed by Brazilian adults. Also, the estimated sample for criterion validity analysis was achieved and presented a similar distribution of sex and macro-region of residence in relation to the general Brazilian population. Furthermore, we validated the NovaFFQ against an instrument, the Nova24h (Neri et al. 2023), also designed and validated to assess food consumption based on the degree of food processing, which ensured that misclassification in the Nova system did not hinder the NovaFFQ validation.

Our sample’s elevated level of schooling is a characteristic of the NutriNet-Brasil study (Santos et al. 2023). This may have facilitated participants’ response, as NovaFFQ has a high cognitive demand. However, it could reduce the external validity of the results, given that only half of the Brazilian population currently completes high school (Brazilian Institute of Geography and Statistics 2019). On other hand, to minimize this issue, we drew the sample stratified by level of education, which allowed us to reach around 20% of the sample with schooling lower than completed college/university. Additionally, the intended sample size for the reproducibility analysis was not reached for some specific socio demographic groups. However, this may not invalidate the results, since another author suggests that a sample between 100 and 200 people is enough for validation studies (Willet 1998).

In conclusion, the NovaFFQ emerges as a valuable instrument that can immediately provide estimates of energy contribution from the Nova food groups for the whole Brazilian population. It is an instrument understood by the population of interest, which presents excellent reproducibility and moderate to substantial criterion validity to evaluate usual food consumption based on the degree of processing.

## Data Availability

All data produced in the present study are available upon reasonable request to the authors.

## Acknowledgements

We would like to thank the Coordination of Superior Level Staff Improvement (grant #88887.619685/2021-00) and the São Paulo Research Foundation (grant #2022/04118-5 and #2021/10993-3), for the financial support provided.

