## Supplementary Material for "A novel food frequency questionnaire for Brazilian adults based on the Nova classification system: development, reproducibility and validation"

Supplementary material 1. NovaFFQ in English (free translation).

**Participant,**

Here you will answer about your usual diet over the last twelve months.
For each listed item, answer by selecting:

1) your frequency of consumption (ranging from “never or <1 time per month” to "daily").
2) your typical portion size consumed using the reference portion available for each item (ranging from “0.5” to “>3,5” portions).

For example, if you usually consume two units of bread per day, select the frequency "daily" and the option "two" for the portion size, knowing that the reference portion it is "1 unit ".

For items that contain more than one food (example: melon or watermelon), record the sum of how often you eat both fruits and the portion size you usually eat.

In the final section, you must record the consumption of items such as sugar, olive oil and ketchup added to drinks, salads, or pizza.

Let's start?

| How often have you eaten the following during the past 12 months? Select **only one** frequency per item. | | | | | | | | | | | How much did you eat each per day (considering the sum of all meals of a day)? Indicate **only one** portion per item. If the response to frequency is "Never or rarely" no need to mark answer for that question. | | | | | | | |
|---|---|---|---|---|---|---|---|---|---|---|---|---|---|---|---|---|---|---|
| Food | Never or rarely | 1 time per month | 2-3 times per month | 1 time a week | 2 times a week | 3 times a week | 4 times a week | 5 times a week | 6 times a week | Daily | Reference portion | 0.5 | 1.0 | 1.5 | 2.0 | 2.5 | 3.0 | +3.5 |
| **1. CEREALS, PASTA, AND PIZZA** | | | | | | | | | | | | | | | | | | |
| Corn | | | | | | | | | | | 1 cob | | | | | | | |
| Canned corn | | | | | | | | | | | 1 tablespoon | | | | | | | |
| Cornflour or northeastern Brazilian couscous | | | | | | | | | | | 1 serving spoon | | | | | | | |
| Homemade popcorn | | | | | | | | | | | 1 bowl average full | | | | | | | |
| Microwave popcorn | | | | | | | | | | | 1 package | | | | | | | |
| Rice | | | | | | | | | | | 1 serving spoon | | | | | | | |
| Homemade or artisanal lasagna, ravioli, cannelloni or other homemade or artisanal stuffed pasta | | | | | | | | | | | 1 piece or 2 servings spoons | | | | | | | |

| | | | | | | | | | | | |
|---|---|---|---|---|---|---|---|---|---|---|---|
| Ready-to-heat pasta dishes (including stuffed pasta and lasagne) | | | | | | | | | | | 1 piece medium or 2 serving spoons |
| Fresh pasta or gnocchi | | | | | | | | | | | 1 serving spoon |
| Instant noodles | | | | | | | | | | | 1 pack |
| Ready-to-heat pizza with brand like Sadia, Perdigão, or pizza from fast-food chains such as Pizza Hut, or Domino's | | | | | | | | | | | 1 slice |
| Homemade or artisanal pizza | | | | | | | | | | | 1 slice |
| **2. BEANS AND LEGUMES** | | | | | | | | | | | |
| Beans | | | | | | | | | | | 1 ladle |
| Fresh pea, chickpea, or lentil | | | | | | | | | | | 1 tablespoon |

| | | | | | | | | | | | |
|---|---|---|---|---|---|---|---|---|---|---|---|
| Canned pea, or chickpea | | | | | | | | | | | 1 tablespoon |
| **3. BURGERS, MEAT AND EGGS** | | | | | | | | | | | |
| Hamburger, cheeseburger, hot dog or other fast-food snack | | | | | | | | | | | 1 unit average |
| Steak burger, chicken or fish nuggets or sticks | | | | | | | | | | | 1 unit of steak burger or 6 units of nuggets |
| Sausage, pepperoni, or frankfurter | | | | | | | | | | | 3 slices of sausage or pepperoni or 1 average frankfurter |
| Dried beef or jerky | | | | | | | | | | | 1 average piece |
| Fresh beef | | | | | | | | | | | 3 medium pieces or 1 steak or serving spoon |

| | | | | | | | | | | | |
|---|---|---|---|---|---|---|---|---|---|---|---|
| Fresh pork | | | | | | | | | | | 1 average slice or 3 ribs |
| Fresh chicken or other poultry | | | | | | | | | | | 1 medium thigh or 1 fillet or 2 medium drumsticks |
| Liver, tongue, gizzard, or other offal | | | | | | | | | | | 1 average piece |
| Egg | | | | | | | | | | | 1 unit |
| Tuna, sardines, or other fish in tin | | | | | | | | | | | 1 tablespoon |
| Cod or other salty fish | | | | | | | | | | | 1 average piece |
| Tilapia, hake, or other fresh fish | | | | | | | | | | | 1 average piece |
| **4. VEGETABLES** | | | | | | | | | | | |
| Lettuce, chard, cress or rocket | | | | | | | | | | | 1 average leaf |

| | | | | | | | | | | | |
|---|---|---|---|---|---|---|---|---|---|---|---|
| Cabbage, kale , or spinach | | | | | | | | | | | 1 tablespoon |
| Endive, Chicory, or escarole | | | | | | | | | | | 1 ladle |
| Tomato or onion | | | | | | | | | | | 1 tomato slice or 2 onion slices |
| Pumpkin, zucchini, or eggplant | | | | | | | | | | | 1 serving spoon |
| Carrot or beet | | | | | | | | | | | 1 tablespoon |
| Broccoli or cauliflower | | | | | | | | | | | 1 average floret |
| Chayote, okra, or green beans | | | | | | | | | | | 1 tablespoon |
| Other vegetables (cucumber, etc) | | | | | | | | | | | 1 tablespoon |
| **5. ROOTS AND TUBERS** | | | | | | | | | | | |
| Cassava | | | | | | | | | | | 1 average piece |
| Sweet potato | | | | | | | | | | | 1 average piece |

| | | | | | | | | | | | |
|---|---|---|---|---|---|---|---|---|---|---|---|
| French fries frozen or purchased in fast-food stores | | | | | | | | | | | 1 average serving or 1/2 dish |
| Potato | | | | | | | | | | | 1 small unit |
| Cassava flour, homemade *"farofa"* | | | | | | | | | | | 1 tablespoon |
| Ready-to-eat *"farofa"* | | | | | | | | | | | 1 tablespoon |
| **6. FRUITS** | | | | | | | | | | | |
| Banana | | | | | | | | | | | 1 unit average |
| Orange, or tangerine | | | | | | | | | | | 1 unit average |
| Apple or pear | | | | | | | | | | | 1 unit average |
| Papaya | | | | | | | | | | | 1 slice average |
| Mango | | | | | | | | | | | 1 unit average |

| | | | | | | | | | | | |
|---|---|---|---|---|---|---|---|---|---|---|---|
| Watermelon or melon | | | | | | | | | | | 1 slice average of watermelon or 2 slice average of melon |
| Others fruits (grape, pineapple, etc) | | | | | | | | | | | 1 slice average |
| **7.CAKES, CANDY, DESSERTS AND BREAKFAST CEREALS** | | | | | | | | | | | |
| Fruit in syrup, or candy made from fruit (e.g. *cocada*, g*oiabada*) | | | | | | | | | | | 1 unit average or 1 tablespoon |
| Candy made from peanut (*paçoca*, *pé-de-moleque*) | | | | | | | | | | | 1 unit average |
| Milk caramel, or hazelnut butter | | | | | | | | | | | 1 tablespoon |
| Homemade flan, coconut pudding, mousse, rice pudding, trifle made from the ingredients | | | | | | | | | | | 1 piece average or 1 serving spoon |

| | | | | | | | | | | | |
|---|---|---|---|---|---|---|---|---|---|---|---|
| Ready-to-eat or powdered flan, coconut pudding, mousse, rice pudding, or trifle | | | | | | | | | | | 1 piece average or 1 serving spoon |
| Porridge, hominy porridge, or corn pudding. | | | | | | | | | | | 1 dish |
| Gelatin | | | | | | | | | | | 1 bowl average |
| Chocolate, or truffle | | | | | | | | | | | 1 unit average |
| Ice cream | | | | | | | | | | | 1 serving |

| | | | | | | | | | | | |
|---|---|---|---|---|---|---|---|---|---|---|---|
| Cookie with or without filling | | | | | | | | | | | 2 units of cookie without filling or 1 unit of cookie stuffed |
| Homemade cake | | | | | | | | | | | 1 slice average |
| Ready-to-eat or powdered cake | | | | | | | | | | | 1 unit small or 1 slice average |
| Breakfast cereals or granola | | | | | | | | | | | 1 bowl of breakfast cereal or 3 tablespoons of granola |
| **8.BREADS AND CRACKERS** | | | | | | | | | | | |
| Homemade bread | | | | | | | | | | | 1 slice average |
| Freshly made unpackaged bread (homemade or artisanal) | | | | | | | | | | | 1 unit average or 2 slices |

| | | | | | | | | | | | |
|---|---|---|---|---|---|---|---|---|---|---|---|
| Sandwich bread | | | | | | | | | | | 1 slice average |
| Others packaged breads | | | | | | | | | | | 1 unit average |
| *Tapioca* | | | | | | | | | | | 1 unit average |
| Homemade or bakery Brazilian cheese bread, savory fried or baked, or pie | | | | | | | | | | | 1 unit average |
| Frozen or packaged Brazilian cheese bread, savory fried or baked, or pie | | | | | | | | | | | 1 unit average |
| Crackers, packaged toast | | | | | | | | | | | 1 unit average |
| Snacks | | | | | | | | | | | 1 pack average |
| **9. PROCESSED MEATS AND CHEESE** | | | | | | | | | | | |

| | | | | | | | | | | | |
|---|---|---|---|---|---|---|---|---|---|---|---|
| Ham, mortadella, or salami | | | | | | | | | | | 1 slice average |
| Mozzarella cheese, or other cheese | | | | | | | | | | | 1 slice average |
| **10. DRINKS** | | | | | | | | | | | |
| Milk | | | | | | | | | | | 1 glass |
| Flavoured yoghurt or ready-to-drink chocolate milk | | | | | | | | | | | 1 glass |
| Milk with chocolate powder | | | | | | | | | | | 1 glass |
| Fresh or pasteurized plain yoghurt | | | | | | | | | | | 1 glass |
| Coffee | | | | | | | | | | | 1 cup |
| Milk with coffee | | | | | | | | | | | 1 cup |
| Fresh 100% fruit juice | | | | | | | | | | | 1 glass |
| Soft drinks (e.g. iced tea, fruit juice) or energy drinks | | | | | | | | | | | 1 glass |

| | | | | | | | | | | | |
|---|---|---|---|---|---|---|---|---|---|---|---|
| Tea and herbal infusions | | | | | | | | | | | 1 cup |
| Soda | | | | | | | | | | | 1 glass |
| Beer | | | | | | | | | | | 1 can or 1 glass |
| Wine | | | | | | | | | | | 1 glass |
| Vodka, whiskey or others drinks alcoholic distilled | | | | | | | | | | | 1 shot |
| **11. NUTS** | | | | | | | | | | | |
| Packaged peanut with brand | | | | | | | | | | | 1 pack small |
| Peanut, nuts with salt or sugar | | | | | | | | | | | 1 serving average |
| Peanut, nuts without salt or sugar | | | | | | | | | | | 1 serving average |
| How often you added those items to ready foods in the last twelve months? (Example: sugar at the coffee, oil at salad, margarine at the bread) Select **only one** option per item. | | | | | | | | | | | How much did you added to foods per day (considering the sum of all meals of a day)? Indicate **only one** portion per item. If the response to frequency is "Never or rarely" no need to mark answer for that question. |
| **12. ITEMS ADDED TO READY DISHES** | | | | | | | | | | | |
| Sugar | | | | | | | | | | | 1 teaspoon |
| Sweetener | | | | | | | | | | | 3 drops or 1 sachet |
| Olive oil or oil | | | | | | | | | | | 1 tablespoon |

| | | | | | | | | | | | |
|---|---|---|---|---|---|---|---|---|---|---|---|
| Butter | | | | | | | | | | | 1 knife tip |
| Margarine | | | | | | | | | | | 1 knife tip |
| Ketchup, mustard, mayonnaise, soy sauce or other ready-made sauces | | | | | | | | | | | 1 tablespoon |
| Cottage cheese or cream cheese | | | | | | | | | | | 1 knife tip |
| Jam | | | | | | | | | | | 1 knife tip |
| Packaged grated cheese | | | | | | | | | | | 1 tablespoon |
| Freshly grated parmesan cheese | | | | | | | | | | | 1 tablespoon |

Supplementary material 2. Table S1. Dietary contribution (% of total energy intake) of Nova subgroups using the Nova24h and the first Nova Food Frequency Questionnaire. Criterion validation study. (n=377)

| Nova groups and subgroups | % of total energy intake | | | | Mean difference | ICC[2] | 95% IC | |
|---|---|---|---|---|---|---|---|---|
| | Nova24h | | NovaFFQ | | | | | |
| | Mean | SE | Mean | SE | | | | |
| **Unprocessed or minimally processed foods** | **50.3** | **0.7** | **56.2** | **0.6** | **6.0** | **0.65** | **0.48** | **0.76** |
| Fruit | 7.8 | 0.3 | 10.7 | 0.4 | 2.9 | 0.69 | 0.54 | 0.78 |
| Red meat | 7.3 | 0.4 | 8.5 | 0.4 | 1.2 | 0.60 | 0.52 | 0.68 |
| Milk and plain yoghurt | 3.6 | 0.2 | 4.8 | 0.3 | 1.1 | 0.65 | 0.56 | 0.71 |
| Eggs | 3.2 | 0.2 | 4.3 | 0.2 | 1.1 | 0.73 | 0.65 | 0.79 |
| Poultry | 3.3 | 0.2 | 4.0 | 0.2 | 0.7 | 0.39 | 0.26 | 0.50 |
| Legumes | 4.5 | 0.2 | 5.0 | 0.2 | 0.5 | 0.62 | 0.53 | 0.69 |
| Pasta | 2.1 | 0.2 | 2.6 | 0.1 | 0.5 | 0.35 | 0.21 | 0.47 |
| Roots and tubers | 1.8 | 0.1 | 2.1 | 0.1 | 0.3 | 0.49 | 0.37 | 0.58 |
| Homemade pies, pastries, pizza | 1.0 | 0.2 | 1.1 | 0.1 | 0.1 | 0.05 | -0.17 | 0.22 |
| Coffee and tea | 0.6 | 0.0 | 0.7 | 0.0 | 0.0 | 0.71 | 0.65 | 0.76 |
| Vegetables | 1.7 | 0.1 | 1.6 | 0.0 | -0.1 | 0.52 | 0.41 | 0.61 |
| Flour | 1.2 | 0.1 | 0.9 | 0.1 | -0.2 | 0.59 | 0.49 | 0.66 |
| Grains | 5.7 | 0.2 | 5.4 | 0.2 | -0.2 | 0.59 | 0.49 | 0.66 |
| Freshly squeezed fruit juice | 2.0 | 0.1 | 1.7 | 0.1 | -0.3 | 0.59 | 0.49 | 0.66 |
| Cassava flour | 1.1 | 0.1 | 0.7 | 0.1 | -0.4 | 0.75 | 0.69 | 0.80 |
| Nuts and seeds without salt, sugar, or oil | 1.6 | 0.2 | 1.2 | 0.1 | -0.5 | 0.55 | 0.45 | 0.64 |
| Fish and seafood | 1.6 | 0.2 | 1.0 | 0.1 | -0.6 | 0.26 | 0.10 | 0.40 |
| Other unprocessed or minimally processed foods | 0.2 | 0.0 | 0.0 | 0.0 | -0.2 | 0.14 | -0.05 | 0.30 |
| **Processed culinary ingredients** | **11.3** | **0.3** | **11.7** | **0.3** | **0.3** | **0.63** | **0.54** | **0.70** |
| Plant oils | 4.8 | 0.1 | 5.9 | 0.1 | 1.1 | 0.55 | 0.38 | 0.66 |
| Animal fats | 1.8 | 0.1 | 1.6 | 0.1 | -0.2 | 0.58 | 0.48 | 0.65 |
| Sugar | 3.1 | 0.2 | 1.9 | 0.1 | -1.3 | 0.47 | 0.30 | 0.59 |
| Other processed culinary ingredients | 1.6 | 0.1 | 2.3 | 0.1 | 0.7 | 0.60 | 0.49 | 0.69 |
| **Processed foods** | **18.5** | **0.6** | **16.7** | **0.5** | **-1.9** | **0.61** | **0.52** | **0.68** |
| Ham and other salted, smoked, or canned meat or fish | 0.7 | 0.1 | 1.8 | 0.1 | 1.1 | 0.26 | 0.09 | 0.40 |
| Wine and beer | 3.6 | 0.3 | 4.0 | 0.3 | 0.4 | 0.73 | 0.66 | 0.78 |
| Canned vegetables and legumes | 0.1 | 0.0 | 0.1 | 0.0 | 0.0 | 0.08 | -0.13 | 0.25 |
| Processed bread | 3.5 | 0.2 | 3.4 | 0.2 | 0.0 | 0.54 | 0.43 | 0.62 |
| Dried and canned fruit | 0.2 | 0.1 | 0.1 | 0.0 | -0.1 | 0.19 | 0.00 | 0.34 |
| Nuts and seeds with salt or sugar | 0.6 | 0.1 | 0.5 | 0.1 | -0.1 | 0.36 | 0.22 | 0.48 |
| Processed cake | 1.2 | 0.2 | 0.8 | 0.1 | -0.4 | 0.07 | -0.14 | 0.24 |
| Processed desserts | 0.5 | 0.1 | 0.1 | 0.0 | -0.4 | -0.03 | -0.25 | 0.15 |

| | | | | | | | | |
|---|---|---|---|---|---|---|---|---|
| Savoury snacks, including croquettes, pastries, and mini pies | 1.2 | 0.2 | 0.7 | 0.1 | -0.5 | 0.12 | -0.07 | 0.28 |
| Cheese | 7.0 | 0.3 | 5.1 | 0.2 | -1.9 | 0.55 | 0.42 | 0.64 |
| **Ultra-processed foods** | **19.8** | **0.6** | **15.4** | **0.5** | **-4.4** | **0.61** | **0.47** | **0.71** |
| Breakfast cereals | 0.4 | 0.1 | 0.6 | 0.1 | 0.1 | 0.69 | 0.62 | 0.75 |
| Bread | 2.5 | 0.2 | 2.6 | 0.2 | 0.1 | 0.55 | 0.44 | 0.63 |
| Other sweetened beverages (fruit juices, energy drinks) | 0.5 | 0.1 | 0.5 | 0.1 | 0.0 | 0.57 | 0.47 | 0.65 |
| Soda | 0.8 | 0.1 | 0.9 | 0.1 | 0.0 | 0.62 | 0.53 | 0.69 |
| Pre-cooked French fries | 0.3 | 0.1 | 0.2 | 0.0 | 0.0 | -0.02 | -0.24 | 0.17 |
| Sauces, dressings, and gravies | 0.6 | 0.1 | 0.5 | 0.0 | 0.0 | 0.25 | 0.08 | 0.38 |
| Instant and canned soups | 0.2 | 0.1 | 0.2 | 0.0 | 0.0 | 0.07 | -0.14 | 0.24 |
| Cream cheese | 0.3 | 0.0 | 0.3 | 0.0 | 0.0 | 0.55 | 0.45 | 0.63 |
| Desserts | 0.9 | 0.1 | 0.8 | 0.1 | -0.1 | 0.29 | 0.13 | 0.42 |
| Milk-based drinks and flavoured yoghurts | 0.9 | 0.1 | 0.8 | 0.1 | -0.1 | 0.55 | 0.44 | 0.63 |
| Margarine | 0.4 | 0.1 | 0.2 | 0.0 | -0.1 | 0.59 | 0.50 | 0.66 |
| Ready-to-heat or ready-to-eat meals | 0.5 | 0.1 | 0.2 | 0.0 | -0.3 | 0.23 | 0.06 | 0.37 |
| Reconstituted meat or fish products | 2.9 | 0.2 | 2.6 | 0.1 | -0.3 | 0.44 | 0.31 | 0.54 |
| Ready-to-heat pizza and pies | 0.8 | 0.1 | 0.4 | 0.1 | -0.4 | 0.02 | -0.19 | 0.20 |
| Ice cream | 1.1 | 0.1 | 0.6 | 0.1 | -0.4 | 0.28 | 0.12 | 0.41 |
| Vodka, tequila, and other distilled alcoholic drinks | 0.7 | 0.1 | 0.3 | 0.1 | -0.5 | 0.40 | 0.27 | 0.51 |
| Crackers | 1.6 | 0.2 | 1.1 | 0.1 | -0.5 | 0.35 | 0.21 | 0.47 |
| Sweets | 2.3 | 0.2 | 1.6 | 0.1 | -0.7 | 0.36 | 0.22 | 0.48 |
| Cakes, cookies, and pies | 1.4 | 0.2 | 0.8 | 0.1 | -0.7 | 0.29 | 0.13 | 0.42 |
| Other ultra-processed foods | 0.7 | 0.1 | 0.2 | 0.0 | -0.5 | 0.19 | 0.01 | 0.34 |

[1] Absolute difference between the first NovaFFQ and the Nova24h

[2] Intraclass correlation coefficients
